# A Preliminary Study on Rapid Quantitative and Qualitative Detection Methods for Apolipoprotein E4 in Plasma

**DOI:** 10.64898/2026.06.24.26356196

**Authors:** Heren Xiao, Ting Wang, Qing Yang, Xiaotong Chen, Yaping Li, Xia Li, Yong Lin, Ben J. Gu

## Abstract

**Objective:** The *APOE* ε4 genotype is the known most significant risk factor for sporadic Alzheimer’s disease (AD), but the association between the blood levels of its encoded apolipoprotein E4 (ApoE4) and AD is poorly understood.

**Methods:** We developed a chemiluminescent quantitative assay and a colloidal gold lateral chromatography qualitative assay to detect ApoE4 protein in plasma samples and compared the results with conventional genotyping.

**Results:** In 185 samples, the plasma ApoE4 protein levels in non-ε4 carriers (ε2/ε3 and ε3/ε3) were 0.07 +/-0.23 and 0.05 +/-0.17 μg /mL, respectively; while the levels in ε4 carriers (ε2/ε4, ε3/ε4, and ε4/ε4) were 4.58 +/-1.96, 4.28 +/-3.49 and 8.27 +/-9.04 μg/mL, respectively. Using a threshold of 1.1 μg /mL, the quantitative method achieved a sensitivity of 100% and a specificity of 99.2%, while the qualitative method showed a concordance rate of 96.2% with genotyping. We also observed that among 133 non-ε4 carriers, one had plasma ApoE4 protein levels higher than the threshold, which would require further investigation.

**Conclusion:** Quantitative and qualitative detection of ApoE4 protein provides as rapid and accurate method for identifying *APOE* ε4 genotype carriers, offering valuable insights for Alzheimer’s disease risk assessment and early intervention.

## 1. Introduction

Alzheimer’s disease (AD) is the most common neurodegenerative disease, characterized by progressive cognitive decline. The number of people with AD is increasing year by year. It is estimated that by 2050, there will be 139 million people with dementia worldwide ^[1]^. As of 2021, the total number of people with Alzheimer’s disease and related dementia in China reached about 16.99 million, an increase of 249.1% compared with 1990 ^[2]^. As China’s population ages, the mortality rate of AD is increasing year by year. The cause of death has risen from 19th in 1990 to 6th in 2019, becoming the sixth leading cause of death^[3]^. AD has seriously affected the health of population.

The *APOE* gene has three main alleles: ε2, ε3 and ε4, corresponding to the proteins ApoE2, ApoE3, and ApoE4, respectively. ApoE2 has a protective effect, ApoE3 is the most common type, while ApoE4 significantly increases the risk of sporadic AD. Individuals homozygous for *APOE* ε4 begin to experience AD-related symptoms at 65.6 years of age, mild cognitive impairment (MCI) at 71.8 years of age, develop dementia at 73.6 years of age, and start to die at 77.2 years of age, 7-10 years earlier than those *APOE* ε3 homozygous carriers ^[4,5]^ .

ApoE4 is a 34.1 kD glycoprotein that is involved in almost all pathological changes in AD, such as promoting Aβ deposition and clearance in the human body, producing neurotoxic fragments, stimulating Tau protein phosphorylation, and impairing mitochondrial function ^[6]^. The driving role of ApoE4 in AD progression can be summarized as lipid metabolism disorder, neurovascular inflammation, and neuronal dysfunction. These interconnected mechanisms help explain why treatments that target Aβ or Tau alone have limited efficacy, especially in *APOE* ε4 carriers ^[7]^. Among the many genetic factors that affect the risk of AD, *APOE* ε4 is currently recognized as the strongest genetic risk factor ^[8-10]^ .

Traditionally, the detection of *APOE* ε4 is mainly carried out by polymerase chain reaction (PCR) and next-generation sequencing (NGS) to detect DNA in peripheral blood, saliva or oral swab samples, and then to determine the individual’s *APOE* genotype ^[8]^. However, in recent years, the direct detection of ApoE4 protein level in blood by immunological methods to infer *APOE* genotype has attracted much attention due to its convenient operation and the fact that it does not require DNA extraction ^[11-15]^ . This study uses colloidal gold test strips and chemiluminescence to detect ApoE4 protein level in blood samples from both qualitative and quantitative perspectives and compares it with traditional genotyping to evaluate its consistency, and then assess whether the detection of ApoE4 protein level in blood can be used as an easy-to-use tool to identify *APOE* ε4 genotype.

## 2. Materials and Methods

### 2.1 Materials

1. Detection kits: Human ApoE4 protein qualitative detection colloidal gold kit (colloidal gold lateral flow immunochromatography) and human ApoE4 protein quantitative detection kit (immuno-chemiluminescence assay). Both kits are independently developed by Qankorey Biotechnology Co., Ltd.
2. Plasma samples: A total of 185 samples were collected, of which 143 were from Jing’an District Central Hospital in Shanghai and 42 were from Shanghai Mental Health Center. This study, as a follow-up to the Ministry of Science and Technology’s major project on proactive health (SQ2018YFC200022), was approved by the Human Ethics Committee of Huashan Hospital, Fudan University (No. 2020-004).

### 2.2 Instruments

The immuno-chemiluminescence analyzer is a product of Yingkai Biotechnology (Shenzhen, China).

### 2.3 Methods

#### 2.3.1 ApoE4 protein detection

1. Following the instructions of the Human ApoE4 Protein Detection Kit (Colloidal Gold Method), qualitative testing was performed on 185 blood/plasma samples. Briefly, the test strip consists of a PVC base with sequentially overlapping sample pad, blood filter pad, conjugate pad, chromatography pad, and absorbent pad. The conjugate pad is coated with 30 nm colloidal gold particles that bind to mouse anti-human ApoE monoclonal antibody. A detection line (T line, coated with rabbit anti-human ApoE4 monoclonal antibody) is located on one side of the conjugate pad, and a control line (C line, coated with goat anti-mouse IgG polyclonal antibody) is located near the absorbent pad. Blood or plasma 10 μL was mixed with 90 μL diluting buffer, added to the sample well. The strip was allowed development for 10 minutes, photos were taken to record and used to interpret negative, weakly positive, and positive results (see additional information for all photos).
2. Following the instructions of the Human ApoE4 Protein Detection Kit (immunochemiluminescence assay, CLIA), quantification was performed on 185 plasma samples. In brief, JSR carboxyl magnetic beads were activated, conjugated with mouse anti-human ApoE monoclonal antibody, and blocked. Rabbit anti-human ApoE4 monoclonal antibody was labeled with acridine ester (AE, type NSP-SA-NHS) and dialyzed and centrifuged. Samples (2 μL each) and 50 μL magnetic beads, 50 μL of the blocking agent (blocking rheumatoid factor interference and reducing non-specific reactions) was mixed and reacted at 37 ^°^C for 5 minutes. The tube was washed, and then 50 μL of AE-labeled antibody was added. The mixture was incubated at 37 ^°^C for 8 minutes, then excitation solution was added and the chemiluminescence intensity was detected at 430 nm.

#### 2.3.2 *APOE* Genotyping

Samples from Shanghai Jing’an District Central Hospital were sent to Hunan Sansure Biotech Co., Ltd (Changsha, China) for genotyping, while 42 plasma samples from Shanghai Mental Health Center have already been genotyped at the Shanghai Mental Health Center but the results were blinded to researchers until ApoE4 measurement was completed.

### 2.4 Statistical Analysis

The experimental results were statistically analyzed using SPSS 18 software.

## 3. Results

### 3.1 Establishment of the standard curve

A standard curve for immunochemiluminescent assay was established, and the experimental results are shown in Figure 1. Within the concentration range of 0-50 μg/mL, there is a significant curve correlation. After piecewise fitting with 5PLC, the curve correlation coefficient r > 0.9900, the main curve correlation coefficient is 0.9973, the calibration curve correlation coefficient is 0.9999, and the deviation of each concentration point is within ±10%.

**Figure 1:**
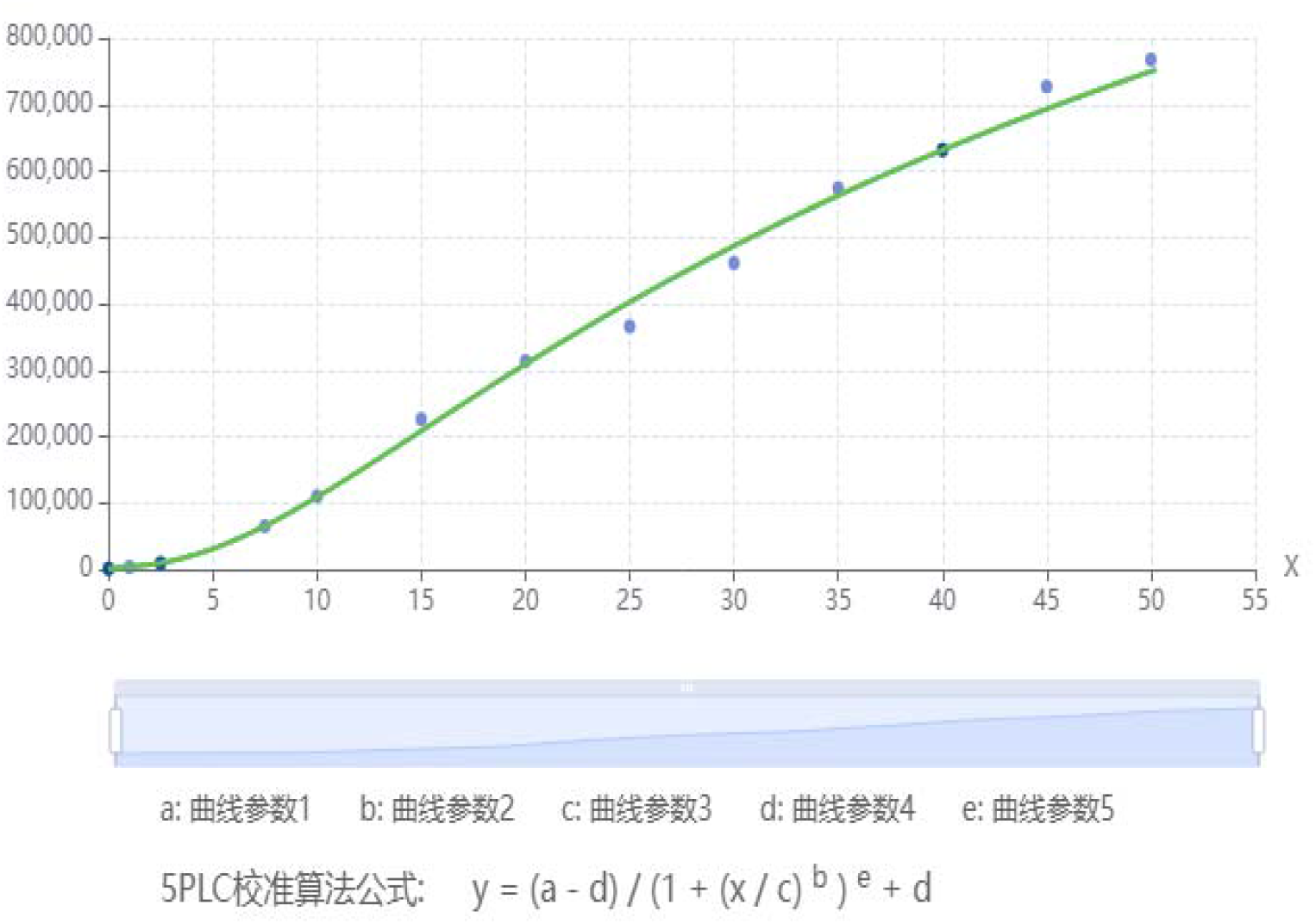
The standard curve of ApoE4 protein quantitation with the immuno-chemiluminescent method.

### 3.2 *APOE* genotyping and ApoE4 protein detection

Genotyping and APOE4 protein detection were performed on 185 plasma samples from Shanghai Jing’an District Central Hospital and Shanghai Mental Health Center . The results are shown in Table 1.

**Table 1a.** Genotype and ApoE4 protein detection results of samples from Jing’an District Central Hospital,Shanghai.

| No. | genotype | Chemiluminescence | Colloidal |
| --- | --- | --- | --- |

|  |  | method (µg/mL) | gold method |
| --- | --- | --- | --- |
| 3 | ε2/ε3 | 1.0 | - / + |
| 4 | ε3/ε3 | 0.6 | - / + |
| 5 | ε2/ε3 | 0.7 | - |
| 6 | ε3/ε3 | 0.1 | - / + |
| 7 | ε3/ε3 | 0.0 | - |
| 8 | ε3/ε3 | 0.0 | - |
| 9 | ε3/ε3 | 0.0 | - |
| 10 | ε3/ε3 | 0.0 | - |
| 11 | ε3/ε3 | 0.0 | - |
| 12 | ε3/ε3 | 0.0 | - |
| 15 | ε3/ε3 | 0.1 | + |
| 16 | ε3/ε4 | 3.7 | + |
| 17 | ε3/ε3 | 0.0 | - |
| 18 | ε3/ε4 | 5.1 | + |
| 19 | ε3/ε3 | 0.0 | - |
| 20 | ε3/ε3 | 0.0 | - / + |
| 21 | ε3/ε3 | 0.0 | - |
| 22 | ε4/ε4 | 18.1 | + |
| 23 | ε3/ε3 | 0.0 | - |
| 24 | ε3/ε3 | 0.0 | - |
| 25 | ε2/ε3 | 0.0 | - |
| 26 | ε3/ε4 | 4.6 | + |
| 27 | ε3/ε3 | 0.0 | - |
| 28 | ε3/ε3 | 0.0 | - |
| 29 | ε2/ε3 | 0.0 | - |
| 30 | ε3/ε3 | 0.0 | - |
| 31 | ε2/ε3 | 0.0 | + |
| 32 | ε3/ε3 | 0.0 | - |
| 33 | ε3/ε3 | 0.4 | + |
| 34 | ε4/ε4 | 23.7 | + |
| 35 | ε2/ε4 | 6.8 | + |
| 36 | ε3/ε3 | 0.0 | - |
| 37 | ε2/ε3 | 0.0 | - |
| 38 | ε3/ε3 | 0.0 | - |
| 39 | ε3/ε4 | 7.4 | + |
| 40 | ε3/ε3 | 0.0 | - / + |
| 41 | ε2/ε3 | 0.0 | - |
| 42 | ε3/ε3 | 0.0 | - |
| 43 | ε3/ε3 | 0.0 | - |
| 44 | ε3/ε4 | 3.6 | + |
| 45 | ε3/ε3 | 0.0 | - |
| 46 | ε3/ε4 | 9.5 | + |
| 47 | $\varepsilon_3/\varepsilon_3$ | 0.0 | - |
| 48 | $\varepsilon_3/\varepsilon_3$ | 0.0 | - |
| 49 | $\varepsilon_3/\varepsilon_3$ | 0.0 | - |
| 50 | $\varepsilon_2/\varepsilon_3$ | 0.0 | - / + |
| 51 | $\varepsilon_2/\varepsilon_3$ | 0.0 | - |
| 52 | $\varepsilon_3/\varepsilon_3$ | 0.4 | - |
| 53 | $\varepsilon_3/\varepsilon_3$ | 0.3 | - |
| 54 | $\varepsilon_3/\varepsilon_3$ | 0.0 | - |
| 55 | $\varepsilon_3/\varepsilon_3$ | 0.0 | - / + |
| 56 | $\varepsilon_3/\varepsilon_3$ | 0.3 | + |
| 57 | $\varepsilon_3/\varepsilon_3$ | 0.0 | - |
| 58 | $\varepsilon_3/\varepsilon_4$ | 21.0 | + |
| 59 | $\varepsilon_2/\varepsilon_3$ | 0.0 | - / + |
| 60 | $\varepsilon_3/\varepsilon_3$ | 0.0 | - |
| 61 | $\varepsilon_3/\varepsilon_3$ | 0.0 | - |
| 62 | $\varepsilon_3/\varepsilon_3$ | 0.0 | - / + |
| 63 | $\varepsilon_3/\varepsilon_3$ | 0.0 | - / + |
| 64 | $\varepsilon_3/\varepsilon_4$ | 6.5 | + |
| 65 | $\varepsilon_3/\varepsilon_3$ | 0.0 | - |
| 66 | $\varepsilon_3/\varepsilon_4$ | 5.3 | + |
| 67 | $\varepsilon_3/\varepsilon_3$ | 0.0 | - |
| 68 | $\varepsilon_2/\varepsilon_3$ | 0.0 | - |
| 69 | $\varepsilon_2/\varepsilon_3$ | 0.0 | - |
| 70 | $\varepsilon_3/\varepsilon_3$ | 0.0 | - |
| 71 | $\varepsilon_3/\varepsilon_3$ | 0.0 | - / + |
| 72 | $\varepsilon_3/\varepsilon_3$ | 0.0 | - |
| 73 | $\varepsilon_3/\varepsilon_3$ | 0.0 | - |
| 74 | $\varepsilon_2/\varepsilon_3$ | 0.0 | - |
| 75 | $\varepsilon_3/\varepsilon_4$ | 3.1 | + |
| 76 | $\varepsilon_2/\varepsilon_3$ | 0.0 | - |
| 77 | **$\varepsilon_3/\varepsilon_3$** | **1.3** | - / + |
| 78 | $\varepsilon_3/\varepsilon_3$ | 0.0 | - |
| 79 | $\varepsilon_4/\varepsilon_4$ | 32.1 | + |
| 80 | $\varepsilon_3/\varepsilon_3$ | 0.0 | - |
| 81 | $\varepsilon_3/\varepsilon_3$ | 0.0 | - |
| 82 | $\varepsilon_2/\varepsilon_3$ | 0.0 | - |
| 83 | $\varepsilon_3/\varepsilon_3$ | 0.0 | - |
| 84 | $\varepsilon_2/\varepsilon_3$ | 0.4 | - / + |
| 85 | $\varepsilon_3/\varepsilon_4$ | 4.6 | + |
| 86 | $\varepsilon_3/\varepsilon_3$ | 0.0 | - |
| 87 | $\varepsilon_3/\varepsilon_3$ | 0.0 | - |
| 88 | $\varepsilon_2/\varepsilon_3$ | 0.0 | - |
| 89 | $\varepsilon_3/\varepsilon_3$ | 0.0 | - |
| 90 | $\varepsilon_3/\varepsilon_4$ | 3.6 | + |
| 91 | $\varepsilon_3/\varepsilon_4$ | 5.4 | + |
| 92 | $\varepsilon_3/\varepsilon_4$ | 5.0 | + |
| 93 | $\varepsilon_3/\varepsilon_3$ | 0.0 | - |
| 94 | $\varepsilon_3/\varepsilon_3$ | 0.0 | + |
| 95 | $\varepsilon_3/\varepsilon_3$ | 0.0 | - |
| 96 | $\varepsilon_4/\varepsilon_4$ | 7.0 | + |
| 97 | $\varepsilon_3/\varepsilon_3$ | 0.0 | - |
| 99 | $\varepsilon_3/\varepsilon_4$ | 4.7 | + |
| 100 | $\varepsilon_2/\varepsilon_3$ | 0.0 | - |
| 101 | $\varepsilon_3/\varepsilon_3$ | 0.0 | - |
| 102 | $\varepsilon_3/\varepsilon_3$ | 0.0 | - |
| 103 | $\varepsilon_3/\varepsilon_3$ | 0.0 | - |
| 104 | $\varepsilon_2/\varepsilon_3$ | 0.0 | - |
| 105 | $\varepsilon_3/\varepsilon_3$ | 0.0 | - |
| 106 | $\varepsilon_2/\varepsilon_3$ | 0.0 | - |
| 107 | $\varepsilon_3/\varepsilon_3$ | 0.0 | - |
| 108 | $\varepsilon_3/\varepsilon_3$ | 0.0 | - |
| 109 | $\varepsilon_3/\varepsilon_3$ | 0.0 | - |
| 110 | $\varepsilon_3/\varepsilon_4$ | 3.9 | + |
| 111 | $\varepsilon_3/\varepsilon_3$ | 0.0 | - |
| 112 | $\varepsilon_2/\varepsilon_3$ | 0.0 | - |
| 113 | $\varepsilon_3/\varepsilon_3$ | 0.0 | - |
| 114 | $\varepsilon_3/\varepsilon_3$ | 0.0 | - |
| 115 | $\varepsilon_3/\varepsilon_3$ | 0.0 | - / + |
| 116 | $\varepsilon_3/\varepsilon_3$ | 0.0 | - |
| 117 | $\varepsilon_3/\varepsilon_4$ | 4.8 | + |
| 118 | $\varepsilon_3/\varepsilon_3$ | 0.0 | - |
| 119 | $\varepsilon_3/\varepsilon_3$ | 0.0 | - |
| 120 | $\varepsilon_3/\varepsilon_3$ | 0.0 | - |
| 122 | $\varepsilon_3/\varepsilon_3$ | 0.0 | - |
| 123 | $\varepsilon_3/\varepsilon_3$ | 0.0 | - |
| 124 | $\varepsilon_2/\varepsilon_3$ | 0.0 | - |
| 125 | $\varepsilon_3/\varepsilon_3$ | 0.0 | - |
| 126 | $\varepsilon_3/\varepsilon_3$ | 0.0 | - |
| 127 | $\varepsilon_3/\varepsilon_3$ | 0.0 | - |
| 128 | $\varepsilon_3/\varepsilon_3$ | 0.0 | - |
| 129 | $\varepsilon_2/\varepsilon_3$ | 0.0 | - |
| 130 | $\varepsilon_3/\varepsilon_3$ | 0.0 | - |
| 131 | $\varepsilon_3/\varepsilon_3$ | 0.0 | - |
| 133 | $\varepsilon_2/\varepsilon_3$ | 0.0 | - |
| 134 | $\varepsilon_2/\varepsilon_3$ | 0.0 | - |
| 135 | $\varepsilon_3/\varepsilon_3$ | 0.0 | - |
| 136 | ε3/ε3 | 0.0 | - |
| 137 | ε3/ε4 | 3.6 | + |
| 138 | ε3/ε3 | 0.1 | - |
| 139 | ε3/ε3 | 0.0 | - |
| 140 | ε2/ε4 | 5.0 | + |
| 141 | ε3/ε3 | 0.0 | - |
| 142 | ε3/ε4 | 4.8 | + |
| 143 | ε3/ε3 | 0.3 | + |
| 144 | ε3/ε4 | 4.2 | + |
| 145 | ε3/ε3 | 0.5 | - / + |
| 146 | ε3/ε3 | 0.0 | - |
| 147 | ε3/ε3 | 0.0 | - |
| 148 | ε3/ε3 | 0.0 | - |
| 149 | ε3/ε3 | 0.0 | - |
| 150 | ε3/ε3 | 0.0 | - / + |

**Table 1b:** Genotype and ApoE4 protein detection results at Shanghai Mental Health Center.

| No. | genotype | Chemiluminescence method (μg/mL) | Colloidal gold method |
| --- | --- | --- | --- |
| LM1229-4 | ε2/ε3 | 0.0 | - |
| LM1884-2 | ε2/ε3 | 0.0 | - |
| LM2025-1 | ε3/ε3 | 0.0 | - |
| LM2394-2 | ε3/ ε4 | 1.6 | + |
| LM2450-1 | ε3/ε4 | 1.7 | + |
| LM2455-2 | ε3/ε3 | 0.0 | - |
| LM2645-1 | ε3/ε4 | 2.4 | + |
| LM2774-3 | ε3/ε3 | 0.0 | - |
| LM2807-1 | ε4/ε4 | 4.3 | + |
| LM2920-1 | ε3/ε3 | 0.0 | - |
| LM2996 | ε4/ε4 | 3.3 | + |
| LM3034-1 | ε4/ε4 | 1.5 | + |
| LM3121-3 | ε3/ε4 | 1.3 | - / + |
| LM3151-3 | ε3/ε4 | 3.6 | + |
| LM3347-1 | ε3/ε4 | 1.8 | + |
| LM3414 | ε4/ε4 | 3.9 | + |
| LM3415 | ε4/ε4 | 4.1 | + |
| LM3416 | ε4/ε4 | 3.4 | + |
| LM3417 | ε4/ε4 | 3.4 | + |
| LM3577 | ε3/ε4 | 4.7 | + |
| LM3615-1 | ε3/ε4 | 1.5 | + |
| LM3736 | ε4/ε4 | 5.9 | + |
| LM3751 | $\epsilon 2/\epsilon 3$ | 0.0 | - |
| LM3847 | $\epsilon 3/\epsilon 3$ | 0.0 | - |
| LM3848 | $\epsilon 4/\epsilon 4$ | 8.5 | + |
| LM3849 | $\epsilon 3/\epsilon 3$ | 0.0 | - |
| LM3856 | $\epsilon 3/\epsilon 3$ | 0.0 | - |
| LM3913 | $\epsilon 3/\epsilon 3$ | 0.0 | - |
| LM3914 | $\epsilon 3/\epsilon 4$ | 2.2 | + |
| LM3920 | $\epsilon 3/\epsilon 3$ | 0.0 | - |
| LM3922 | $\epsilon 3/\epsilon 4$ | 1.1 | + |
| LM3923 | $\epsilon 3/\epsilon 4$ | 1.8 | + |
| LM3948-1 | $\epsilon 4/\epsilon 4$ | 3.0 | + |
| LM3955-1 | $\epsilon 3/\epsilon 4$ | 2.0 | + |
| LM4010 | $\epsilon 2/\epsilon 4$ | 2.0 | + |
| LM4062 | $\epsilon 4/\epsilon 4$ | 1.9 | + |
| LM4069 | $\epsilon 3/\epsilon 4$ | 1.8 | + |
| LM4108 | $\epsilon 3/\epsilon 3$ | 0.0 | - / + |
| LM4117 | $\epsilon 3/\epsilon 3$ | 0.0 | - |
| LM4144 | $\epsilon 3/\epsilon 3$ | 0.0 | - |
| LM-4297 | $\epsilon 3/\epsilon 3$ | 0.0 | - |
| LM474-S1 | $\epsilon 3/\epsilon 4$ | 3.7 | + |

**Table 2.** Genotypes and ApoE4 protein concentrations.

| Genotype | Number | Protein concentration ($\mu\text{g/mL}$) | | | |
| --- | --- | --- | --- | --- | --- |
|  |  | Mean +/- SD | Middle | Min | Max |
| $\epsilon 2/\epsilon 3$ | 28 | 0.07 +/- 0.23 | 0.0 | 0.0 | 1.0 |
| $\epsilon 3/\epsilon 3$ | 105 | 0.05 +/- 0.17 | 0.0 | 0.0 | 1.3 |
| $\epsilon 2/\epsilon 4$ | 3 | 4.58 +/- 1.96 | 5.0 | 2.0 | 6.8 |
| $\epsilon 3/\epsilon 4$ | 34 | 4.28 +/- 3.49 | 3.7 | 1.1 | 21.0 |
| $\epsilon 4/\epsilon 4$ | 15 | 8.27 +/- 9.04 | 4.1 | 1.5 | 32.1 |

#### 3.2.1 Immuno chemiluminescence immunoassay for ApoE4 protein detection

The samples from the two hospitals were combined and statistically analyzed for each genotype and ApoE4 protein concentration, as follows:

##### 3.2.1.1 ApoE4 protein concentration can distinguish ε4 genotype carrier status

Based on genotyping results, 52 individuals were carriers of the *APOE* ε4 genotype, and 133 were non-carriers. ROC curve analysis was used to analyze the differential diagnostic value of ApoE4 protein relative expression level for ε4 genotype carrier status. The results showed an AUC of 0.999(95% CI:0.996-1.000), indicating that ApoE4 protein expression level has a very high diagnostic effect on ε4 genotype carriers (Figure 2). Based on the maximum Youden index, the optimal cutoff value was determined to be 1.12 μg/mL, at which only one ε3/ε3 carrier showed a false positive, with a diagnostic sensitivity of 1.000(95% CI:0.931-1.000, specificity of 0.992(95% CI:0.959-0.999), Youden index of 0.992 and accuracy of 99.5%(95% CI:0.970-0.999). This indicates that using ApoE4 protein expression of 1.12 μg/mL as the diagnostic threshold can effectively distinguish *APOE* ε4 genotype carriers and non-carriers.

**Figure 2.**
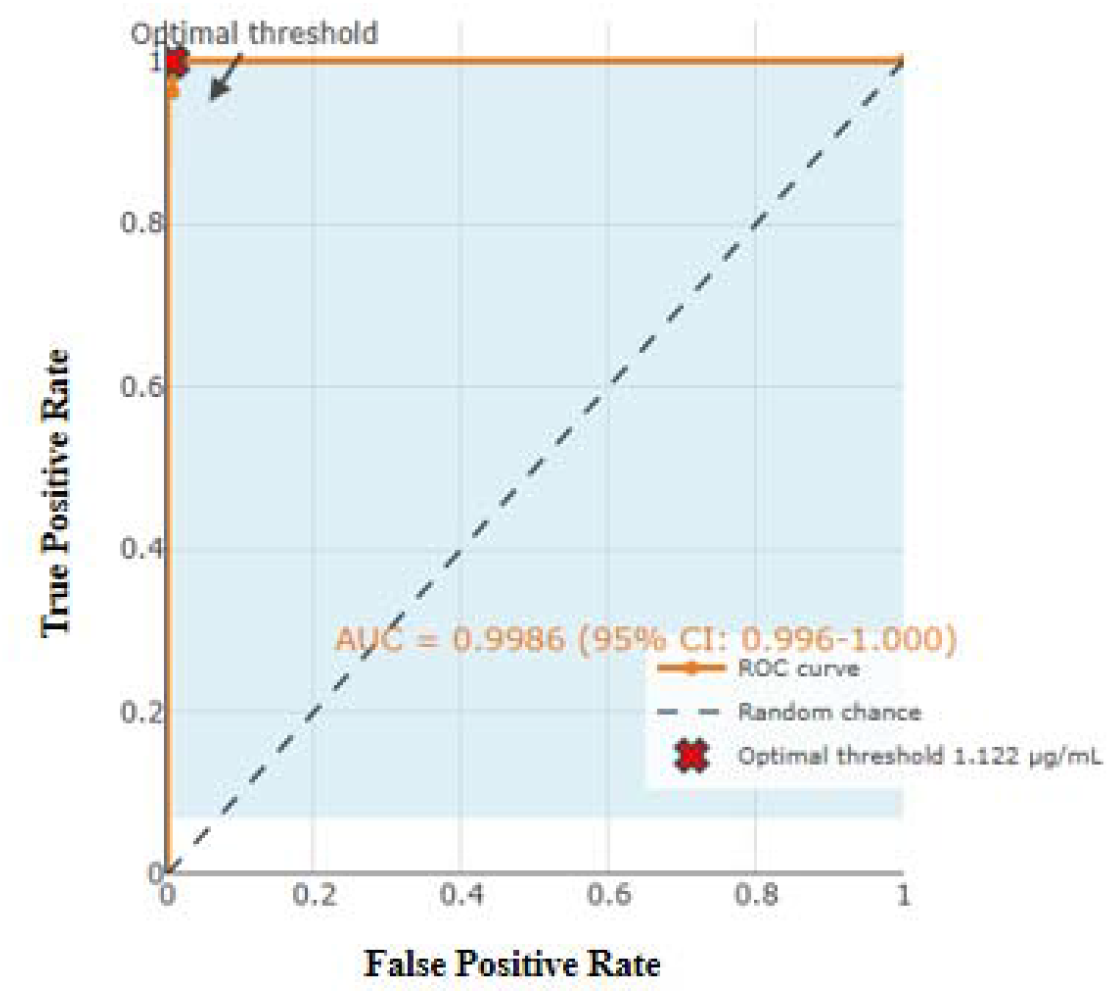
ROC curves showing the differentiation of ε4 genotype carriers by ApoE4 protein concentration.

##### 3.2.1.2 ApoE4 protein concentration is insufficient to distinguish between ε4 heterozygotes and homozygotes

Based on genotyping results, of the 15 *APOE* ε4 homozygotes and 37 heterozygotes, ROC curve analysis showed an AUC of 0.514(95% CI:0.359-0.668), close to the diagonal line, indicating poor distinguishing ability (Figure 3). The optimal threshold was 2.97 μg/mL, with a sensitivity of only 46.7%(95% CI:0.218-0.730), a specificity of 70.3%(95% CI:0.535-0.832), and an accuracy of 63.5% (95% CI:0.492-0.758). Therefore, plasma ApoE4 protein concentration alone is insufficient to effectively distinguish *APOE* ε4 homozygotes and heterozygotes.

**Figure 3.**
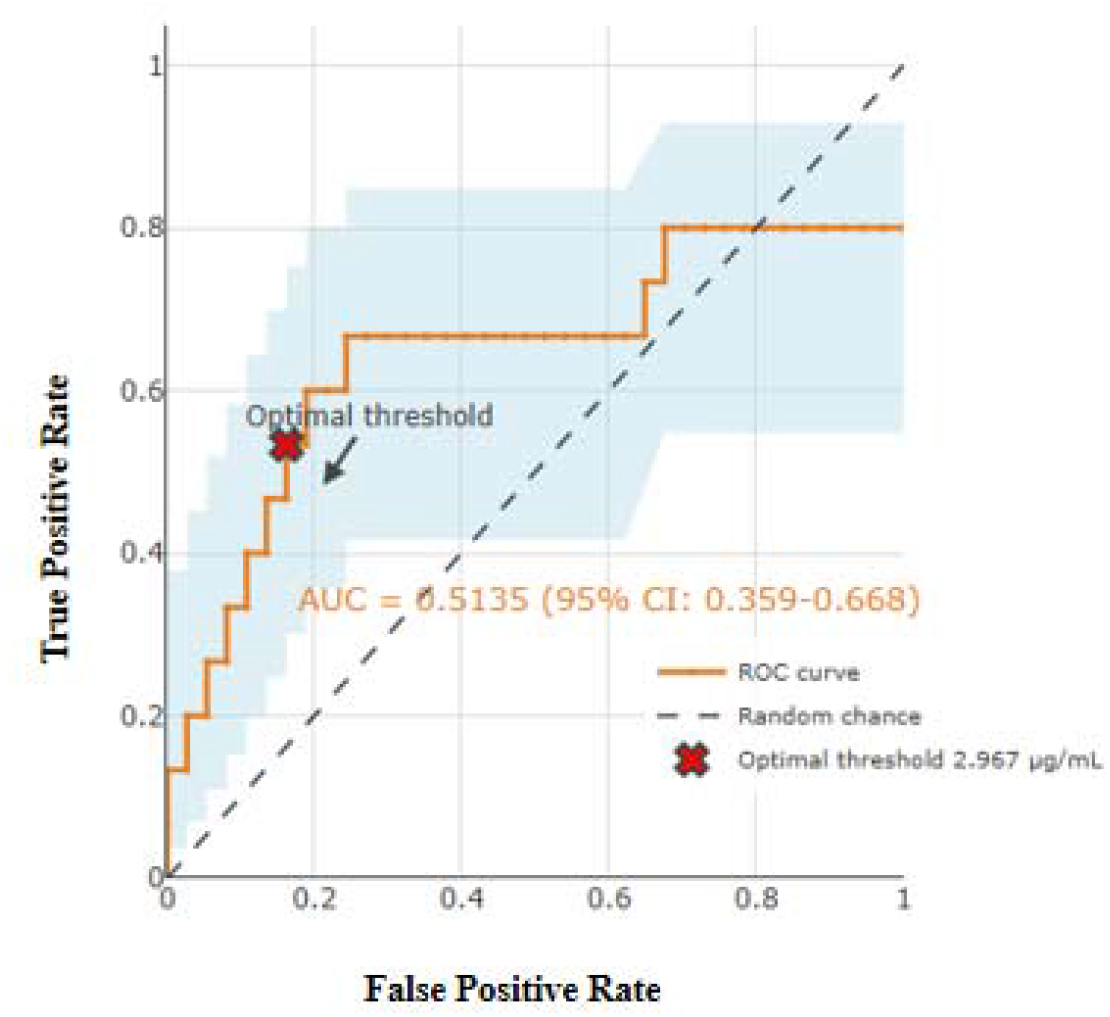
ROC curves showing the differentiation between homozygous and heterozygous ε4 genotypes based on ApoE4 protein concentration.

#### 3.2.2 Detection of ApoE4 protein in plasma using colloidal gold method

We tested samples from two hospitals using colloidal gold test strips, photographed them, and analyzed the positive, weakly positive, and negative results, then compared them with the results of each genotype. The results are shown in Table 1 and summarized in Table 3. Photos of all test strips can be downloaded from https://doi.org/10.6084/m9.figshare.32413287.

**Table 3.** Genotype and ApoE4 colloidal gold test strip results.

| Genotype | Number | Protein Amount |  |  | Consistency |
| --- | --- | --- | --- | --- | --- |
|  |  | Positive | Weak positive | Negative |  |
| $\epsilon 2/\epsilon 3$ | 28 | 1 | 4 | 23 | 96.4% |
| $\epsilon 3/\epsilon 3$ | 10 5 | 5 | 13 | 87 | 95.2% |
| $\epsilon 2/\epsilon 4$ | 3 | 3 | 0 | 0 | 100% |
| $\epsilon 3/\epsilon 4$ | 3 4 | 33 | 1 | 0 | 97.1 % |
| $\epsilon 4/\epsilon 4$ | 15 | 15 | 0 | 0 | 100% |
| Overall compliance rate |  |  |  |  | 96.2% |

Our results show that the colloidal gold method can also effectively distinguish ε4 genotype carriers, with an efficacy rate as high as 96.2%(95% CI:0.924-0.982) if weak positives are considered negative. Furthermore, the convenience and privacy of colloidal gold can be used as an initial screening tool for *APOE* ε4 genotyping in community neighborhood and hospitals.

## 4. Discussion

This study demonstrates that rapid and simple quantitative and qualitative methods can accurately screening for *APOE* ε4 genotype carriers by detecting peripheral blood ApoE4 protein. Similar to previous studies ^[14,15]^, our quantitative analysis also showed significant individual differences in blood ApoE4 protein levels in the population. How these differences arise and whether they are related to the progression of Alzheimer’s disease (AD) is a topic of great interest to researchers. An even more interesting finding is that a small subset of non-*APOE* ε4 genotype carriers had low concentrations of ApoE4 protein in their blood, with some even reaching ε4 heterozygous levels. Whether these ApoE4 proteins are due to methodological interference, such as the formation or breakage of cysteine disulfide bonds, the binding of ApoE3 to different lipid/protein/peptide, or somatic mutations warrants further investigation.

While carriers and non-carriers of the ε4 genotype can be identified by the quantitative methods, it is not possible to effectively distinguish between ε4 heterozygotes and homozygotes. Simultaneous detection of total ApoE protein levels might solve this problem. It has been reported that the ratio of ApoE4 to total ApoE can effectively differentiate between ε4 heterozygotes and homozygotes ^[14,15]^. This method may be more suitable for future clinical and research applications.

Our results show that the use of colloidal gold test strips to detect ApoE4 protein in finger-prick blood can also effectively distinguish between carriers and non-carriers of the ε4 genotype. Although the accuracy (96.2%) is not as good as that of quantitative methods (99.5%), it is faster, more convenient and more private, making it more suitable for self-testing at home and large-scale health screening of the elderly^[16]^. This finger-prick blood ApoE4 protein test strip can also be used as a supplement to our other self-testing-based early screening tool for AD: the urine β-amyloid protein test kit (Qankorey® One-Step Dementia Risk Test Kit), because we found that in cognitively normal individuals, the positive rate of the urine β-amyloid protein test kit for ε4 genotype carriers was significantly lower than that for non-carriers ^[17,18]^ .

Recent large-scale community-based population studies, using blood pTau217 levels, have found that the proportion of elderly individuals exhibiting pathological manifestations of AD increases with age, reaching 3.91%, 2.47%, 7.69%, 18.0%, 28.3%, 44.1%, 57.9%, and 65.2% in the 58-59, 60-64, 65-69, 70-74, 75-79, 80-84, 85-89 and 90+ age groups, respectively ^[19]^, very similar to our urine β-amyloid assay kit results in Chinese communities and among older adults in Australia ^[17,18]^. These results highlight the urgency of early screening in older adults, as nearly 95% of the population is not protected by ApoE2 and is at great risk of developing AD later in life. ^[20]^, and there is currently no clinical treatment specifically targeting on ApoE4. With the popularization of early screening and the improvement of public awareness, there will be more and more solutions for AD in the future.

## 5. Conclusion

Quantitative and qualitative detection of ApoE4 protein levels in blood can effectively and accurately identify APOE ε4 genotype carriers and non-carriers.

## Data Availability

All data produced in the present study are available upon reasonable request to the authors

## 6. Acknowledgments

This research was supported by the grant from Shanghai Fifth People’s Hospital (No. 2022WYZD03) and indirectly supported by the major project of the Ministry of Science and Technology (SQ2018YFC200022 ).

